# Using the Emergency Care Data Set for the epidemiological surveillance of Children and Young People aged less than 18 years: a case study of COVID-19 in England 2020-2023

**DOI:** 10.1101/2024.07.26.24310711

**Authors:** Jacob Boateng, Clarissa Oeser, Giulia Seghezzo, Katie Harman, Gavin Dabrera, Harriet Webster, Russell Hope, Simon Thelwall, Theresa Lamagni

## Abstract

**Background:** The Emergency Care Data Set provides insight into emergency care activity in England, and combined with COVID-19 surveillance data, can provide new insights into acute COVID-19 infection.

**Methods:** This study identified individuals <18 years old who tested positive for SARS-CoV-2 between February 2020 and March 2023 and attended emergency care 1-14 days after a positive test. The study’s main objective was to explore ED attendance outcomes by demographic characteristics.

**Results:** There were significant differences (p < 0.05) across most of the characteristics of <18s admitted to hospital from emergency departments, and those who were discharged from ED. <18s in IMD decile 1 (14.9%) made up the highest proportion of admissions, with those in less deprived areas having a greater proportion of individuals discharged from ED. February to August 2020 (1.5%) and September 2022 to March 2023 (2.8%) saw the highest proportion of <18 cases attending ED, though the highest number of cases were seen between September 2021 and February 2022.

**Conclusions:** There is great value in the use of ECDS. It facilitates quick, regular insights into the health outcomes of key demographics, and provides a window into the health-seeking behaviours of individuals. Furthermore, outcomes of emergency care attendance can potentially inform assessments of infection severity across multiple demographics during outbreaks and pandemics.

## Introduction

Public health surveillance of communicable diseases in England uses diverse data sources to develop a comprehensive understanding of disease trends, capitalising on the strengths of individual sources. For children, monitoring emergency department (ED) attendance is of particular interest as many of these infections may present urgently to acute health services. The Emergency Care Data Set (ECDS) is a key dataset reporting healthcare activity in emergency departments in England, with data being reported by clinicians to capture information about patients attending ED. The purpose of ECDS is to better understand patient capacity and demand while looking to improve patient care and allow for better communication and planning across healthcare services (1).

Linked with laboratory data, these data can provide valuable insights on infections, such as SARS-CoV-2. ECDS forms part of the Hospital Episode Statistics (HES) data product (managed by the Secondary Uses Service (SUS)) and is reported at regular intervals by all organisations providing publicly funded acute NHS health services in England. HES is part of a Commissioning Data Set, and contains information on hospital admissions, outpatient appointments and attendances, and historic A&E attendances (ECDS replaced the HES A&E Commissioning Dataset in 2020), submitted to NHS England (2). In contrast to ECDS, HES data is recorded by clinical coders in hospitals, who translate discharge summary information, written by clinicians, into codes (3,4). The purpose of this analysis is to demonstrate the potential of ECDS to describe infectious disease epidemiology by investigating COVID-19 episodes in children and young people (CYP) attending emergency departments.

## Methods

Episodes of SARS-CoV-2 infection identified through PCR or self-reported lateral flow testing (5) in patients aged less than 18 years at the time of specimen collection were identified from the Second-Generation Surveillance System (SGSS); a system developed by UK Health Security Agency which collects routine infectious disease laboratory surveillance data (6), with specimen collection dates between 3^rd^ February 2020 and 31^st^ March 2023.

These episodes of infection were linked by 2 unique identifiers to the Emergency Care Data Set (ECDS) to identify any relevant records of attendance across all types of ED, including outcomes of attendance: discharge, admission to short stay unit (<24 hr in ED), admission to inpatient care, or transfers to other acute trusts (1). ED attendances were included if occurring 1-14 days after a positive COVID-19 test collected between 3^rd^ February 2020 and 31^st^ March 2023.

Attendances were excluded if they occurred on the same day as the positive test result, to remove the potential of including CYP who were identified by testing in emergency care who may have attended for another reason. They were also excluded if they were flagged with an ICD10 code indicating the main reason for attendance was injury-related, or if no NHS number was available. These episodes were then deduplicated to the person level using a personal identifier to ensure each person was only counted once. The person-level records with linked ECDS entries were merged using NHS number to the National Immunisation Management System (NIMS), a system which records the vaccination information of everyone registered with the NHS, to gather COVID-19 vaccination history (7,8).

Person-level records with linked ECDS entries were described according to sex, age group, ethnicity, index of multiple deprivation 2019 (IMD), geographic region of residence, time period of specimen date and vaccination status. Each time period was split into 6/7-month intervals between February 2020 and March 2023 to reflect the circulation of dominant variants within these timeframes as well as any seasonal variations. Available variant information for sequenced tests was also provided using published SARS-CoV-2 variant assignments (8) (Table 1).

**Table 1.** Descriptive characteristics of total under 18 cases, emergency attendances, and attendance outcomes (admissions/transfers and discharges/short stays), with Chi-Squared test.

| Variables |  | Total number of cases<br>(n = 3,769,882) |  | Total attendances*<br>(n = 26,604) |  | Admissions/<br>transfers<br>(n= 4,563) |  | Discharges/<br>short stays<br>(n= 22,041) |  | χ<sup>2</sup> p-value<br>(admissions/<br>transfers vs<br>discharges/short<br>stays) |
| --- | --- | --- | --- | --- | --- | --- | --- | --- | --- | --- |
|  |  | n | % | n | % | n | % | n | % |  |
| Sex | Female | 1,894,444 | 50.25 | 13,858 | 52.09 | 2,284 | 50.05 | 11,574 | 52.51 | 0.005 |
|  | Male | 1,874,671 | 49.73 | 12,744 | 47.9 | 2,278 | 49.92 | 10,466 | 47.48 |  |
|  | Unknown | 767 | 0.02 | 2 | 0.01 | 1 | 0.02 | 1 | 0.00 |  |
| Age | <1 | 62,501 | 1.66 | 3,851 | 14.48 | 918 | 20.12 | 2,933 | 13.31 | <0.001 |
|  | 1-4 | 331,566 | 8.80 | 4,847 | 18.22 | 870 | 19.07 | 3,977 | 18.04 |  |
|  | 5-9 | 1,081,362 | 28.68 | 5,243 | 19.71 | 714 | 15.65 | 4,529 | 20.55 |  |
|  | 10-14 | 1,522,651 | 40.39 | 7,020 | 26.39 | 1,123 | 24.61 | 5,897 | 26.75 |  |
|  | 15-17 | 771,802 | 20.47 | 5,643 | 21.21 | 938 | 20.56 | 4,705 | 21.35 |  |
| Ethnicity | Asian | 334,740 | 8.88 | 2,954 | 11.10 | 507 | 11.11 | 2,447 | 11.10 | <0.001 |
|  | Black | 99,614 | 2.64 | 870 | 3.27 | 195 | 4.27 | 675 | 3.06 |  |
|  | Mixed | 194,179 | 5.15 | 1,464 | 5.50 | 240 | 5.26 | 1,224 | 5.55 |  |
|  | Other | 44,500 | 1.18 | 536 | 2.01 | 107 | 2.34 | 429 | 1.95 |  |
|  | Unknown | 147,301 | 3.91 | 2,161 | 8.12 | 524 | 11.48 | 1,637 | 7.43 |  |
|  | White | 2,949,548 | 78.24 | 18,619 | 69.99 | 2,990 | 65.53 | 15,629 | 70.91 |  |
| IMD Decile Group† | 1<sup>st</sup> (Most Deprived) | 370,416 | 9.84 | 3,497 | 13.17 | 681 | 14.92 | 2,816 | 12.78 | 0.005 |
|  | 2<sup>nd</sup> | 350,943 | 9.32 | 3,236 | 12.19 | 553 | 12.12 | 2,683 | 12.17 |  |
|  | 3<sup>rd</sup> | 344,956 | 9.16 | 2,943 | 11.09 | 538 | 11.79 | 2,405 | 10.91 |  |
|  | 4<sup>th</sup> | 347,189 | 9.22 | 2,750 | 10.36 | 447 | 9.80 | 2,303 | 10.45 |  |
|  | 5<sup>th</sup> | 357,051 | 9.48 | 2,561 | 9.65 | 419 | 9.18 | 2,142 | 9.72 |  |
|  | 6<sup>th</sup> | 372,869 | 9.90 | 2,479 | 9.34 | 399 | 8.74 | 2,080 | 9.44 |  |
|  | 7<sup>th</sup> | 373,836 | 9.93 | 2,297 | 8.65 | 385 | 8.44 | 1,912 | 8.67 |  |
|  | 8<sup>th</sup> | 392,110 | 10.41 | 2,238 | 8.43 | 382 | 8.37 | 1,856 | 8.42 |  |
|  | 9<sup>th</sup> | 411,202 | 10.92 | 2,258 | 8.51 | 359 | 7.87 | 1,899 | 8.62 |  |
|  | 10<sup>th</sup> (Least deprived) | 445,152 | 11.82 | 2,284 | 8.60 | 387 | 8.48 | 1,897 | 8.61 |  |
| Region† | East Midlands | 328,950 | 8.73 | 2,260 | 8.51 | 377 | 8.26 | 1,883 | 8.54 | <0.001 |
|  | East of England | 489,585 | 12.99 | 3,194 | 12.03 | 488 | 10.69 | 2,706 | 12.28 |  |
|  | London | 473,120 | 12.55 | 4,610 | 17.37 | 735 | 16.11 | 3,875 | 17.58 |  |
|  | North East | 178,164 | 4.73 | 1,726 | 6.50 | 321 | 7.03 | 1,405 | 6.37 |  |
|  | North West | 478,712 | 12.70 | 3,723 | 14.03 | 779 | 17.07 | 2,944 | 13.36 |  |
|  | South East | 669,319 | 17.75 | 3,992 | 15.04 | 624 | 13.68 | 3,368 | 15.28 |  |
|  | South West | 391,274 | 10.38 | 1,917 | 7.22 | 338 | 7.41 | 1,579 | 7.16 |  |
|  | West Midlands | 404,652 | 10.73 | 2,777 | 10.46 | 419 | 9.18 | 2,358 | 10.70 |  |
|  | Yorkshire and Humber | 351,948 | 9.34 | 2,344 | 8.83 | 469 | 10.28 | 1,875 | 8.51 |  |
| Variant | V-22APR-04 (Omicron BA.5) | 3,636 | 0.42 | 152 | 2.11 | 62 | 1.36 | 90 | 0.41 | <0.001 |
|  | VOC-20DEC-01 (Alpha) | 33,017 | 3.85 | 294 | 4.09 | 70 | 1.53 | 224 | 1.02 |  |
|  | VOC-21APR-02 (Delta) | 404,439 | 47.10 | 3,274 | 45.53 | 664 | 14.55 | 2,610 | 11.84 |  |
|  | VOC-21NOV-01 (Omicron BA.1) | 297,124 | 34.60 | 2,234 | 31.07 | 416 | 9.12 | 1,818 | 8.25 |  |
|  | VUI-21OCT-01 (Delta AY4.2) | 32,393 | 3.77 | 185 | 2.57 | 33 | 0.72 | 152 | 0.69 |  |
|  | VUI-22JAN-01 (Omicron BA.2) | 40,176 | 4.68 | 491 | 6.83 | 109 | 2.39 | 382 | 1.73 |  |
|  | Other | 3,997 | 0.47 | 155 | 2.16 | 74 | 1.62 | 81 | 0.37 |  |
|  | Unclassified/Undetermined | 43,880 | 5.11 | 406 | 5.65 | 114 | 2.50 | 292 | 1.32 |  |
|  | Missing/Unknown | 2,911,220 | 77.22 | 19,413 | 72.97 | 3,021 | 66.21 | 16,392 | 74.37 |  |
| Time period | Time Period 1<br>(03/02/2020 - 31/08/2020) | 8,737 | 0.23 | 132 | 0.50 | 42 | 0.92 | 90 | 0.41 | <0.001 |
|  | Time Period 2<br>(01/09/2020 - 28/02/2021) | 338,229 | 8.97 | 2,534 | 9.52 | 529 | 11.59 | 2,005 | 9.10 |  |
|  | Time Period 3<br>(01/03/2021 - 31/08/2021) | 458,583 | 12.16 | 3,987 | 14.99 | 638 | 13.98 | 3,349 | 15.19 |  |
|  | Time Period 4<br>(01/09/2021 - 28/02/2022) | 2,548,789 | 67.61 | 15,071 | 56.65 | 2,299 | 50.38 | 12,772 | 57.95 |  |
|  | Time Period 5<br>(01/03/2022 - 31/08/2022) | 376,177 | 9.98 | 3,784 | 14.22 | 748 | 16.39 | 3036 | 13.77 |  |
|  | Time Period 6<br>(01/09/2022 - 31/03/2023) | 39,367 | 1.04 | 1,096 | 4.12 | 307 | 6.73 | 789 | 3.58 |  |
| Vaccination status | Vaccinated ($\geq 1$ dose) | 1,549,721 | 41.11 | 8,487 | 31.90 | 1,391 | 30.48 | 7,096 | 32.19 | 0.062 |
|  | Unvaccinated | 2,213,700 | 58.72 | 18,086 | 67.98 | 4 | 0.09 | 27 | 0.12 |  |
|  | Unknown/Unlinked | 6,461 | 0.17 | 31 | 0.12 | 3,168 | 69.43 | 14,918 | 67.68 |  |

† 61 ED attendance records contained missing IMD and region data (n = 4,550 admissions/transfers; n = 21,993 discharged/ short stays; n = 26,543 overall attendances); 4,158 cases aged 0-17 contained missing IMD and region data (n = 3,765,724)
\*The 8 CYP who died following attendance have been excluded

## Results

Between 3^rd^ February 2020 and 31^st^ March 2023, 3,769,882 children and young people (0-17y) had a SARS-CoV-2 positive test result. Of those, 26,612 (0.7%) were identified as having attended emergency care. Of the children and young people who attended emergency care, 4,563 (17.1%) were subsequently admitted as inpatients or transferred to other trusts, while 22,041 (82.8%) either were admitted to a short stay assessment unit or were discharged. The remaining 8 (0.03%) individuals died while in emergency care. Table 1 describes the characteristics of those admitted/transferred in comparison to those who had a short stay/were discharged, with the total number of cases and attendances present for reference. Moving forward admitted/transferred will be referred to as “admitted” and short stay/were discharged as “discharged”.

Across age groups, 40.4% of the cohort were between the ages of 10-14, also making up the highest percentage of admissions (24.6%) and discharges (26.8%). Those under 1 year of age showed the largest difference between the emergency care groups, making up 20% of admissions, but only 13% of discharges. There was an even distribution among males and females testing positive in CYP, with females making up 50.3% of the cohort, and males 49.7%. Of those that attended ED, more females were discharged (52.5%) than males (47.5%).

Whilst White CYP comprised 78.2% of COVID-19 cases in the dataset, they made up a comparatively smaller proportion of ED attendances (70.0%). When observing the proportion of admissions following ED attendance, 65.5% were of White ethnicity, compared to 70.9% of those discharged. Following White CYP, those of Asian and Mixed ethnicities made up highest proportion of ED attendances (11.1% and 5.5% respectively). Among the Black and Other ethnicities, admissions to hospital (4.3% - Black, and 2.3% - Other) were higher than discharges (3.1% - Black, and 2.0% - Other). Those of Asian ethnicity meanwhile made up a similar percentage of admissions and discharges (both 11.1%).

In the CYP case cohort, a higher proportion of 0-17-year-olds in the least deprived deciles tested positive for COVID-19, however those in IMD deciles 1 and 2 (the most deprived) accounted for highest proportion of ED attendances (IMD 1 - 13.2%, IMD 2 – 12.2%). In addition, CYP in IMD decile 1 made up the highest proportion of admissions (14.9%) and a higher proportion of those in IMD deciles 1-3 were admitted than discharged, while those in IMD deciles 4-10 had a higher proportion of discharged than admitted.

Regionally, the South East had the highest proportion of COVID-19 cases among CYP, (17.8%), and the North East contained the lowest percentage of CYP cases (4.7%) and CYP attendances to ED (6.5%). Meanwhile, London had the highest proportion of CYP attending emergency care at 17.4% (while having the fourth highest proportion of COVID-19 cases among CYP at 12.6%). Across ED attendance outcomes, the North West had the highest percentage of young people being admitted (17.1%), while London had the highest percentage of discharges (17.6%). Only the North West and North East had a higher proportion of admitted CYP than discharged, all other regions had a higher proportion of discharged. However, it is important to note that the populations in each region differ, so it is difficult to make regional comparisons by just observing the proportions in Table 1. To make more accurate ED attendance comparisons between regions, we would need to adjust for regional population denominators for 0–17-year-olds.

The highest number of cases in the cohort occurred between 1^st^ September 2021 and 28^th^ February 2022 (Time period 4), accounting for 67.6% of positive tests among CYP (Table 1, Figure 1). Likewise, the greatest percentage of ED attendances (56.7%) occurred in Time Period 4, along with admissions (50.4%), as well as discharges (58.0%). This is further supported by the majority of CYP cases with an assigned variant either having the Delta (47.1%) or Omicron BA.1 (34.6%) variants (Table 1, Figure 1). During this period, the dominant variant in circulation in England transitioned from the Delta variant to Omicron BA.1 (9). However, the periods with highest proportion of CYP cases who attended ED were Period 1 (i.e. of all cases in Period 1 (n = 8,737), 1.5% attended ED (n = 132)) and Period 6 (2.8%), as well as having a higher proportion of admittances to hospital following ED attendance, with 32% of attendances being admitted (n = 42) in Period 1, and 28% of attendances (n = 1,096) being admitted (n = 307) in Period 6. Both periods correspond to times with limited access to vaccinations (Period 1) and limited access to community testing (Period 6).

**Figure 1.**
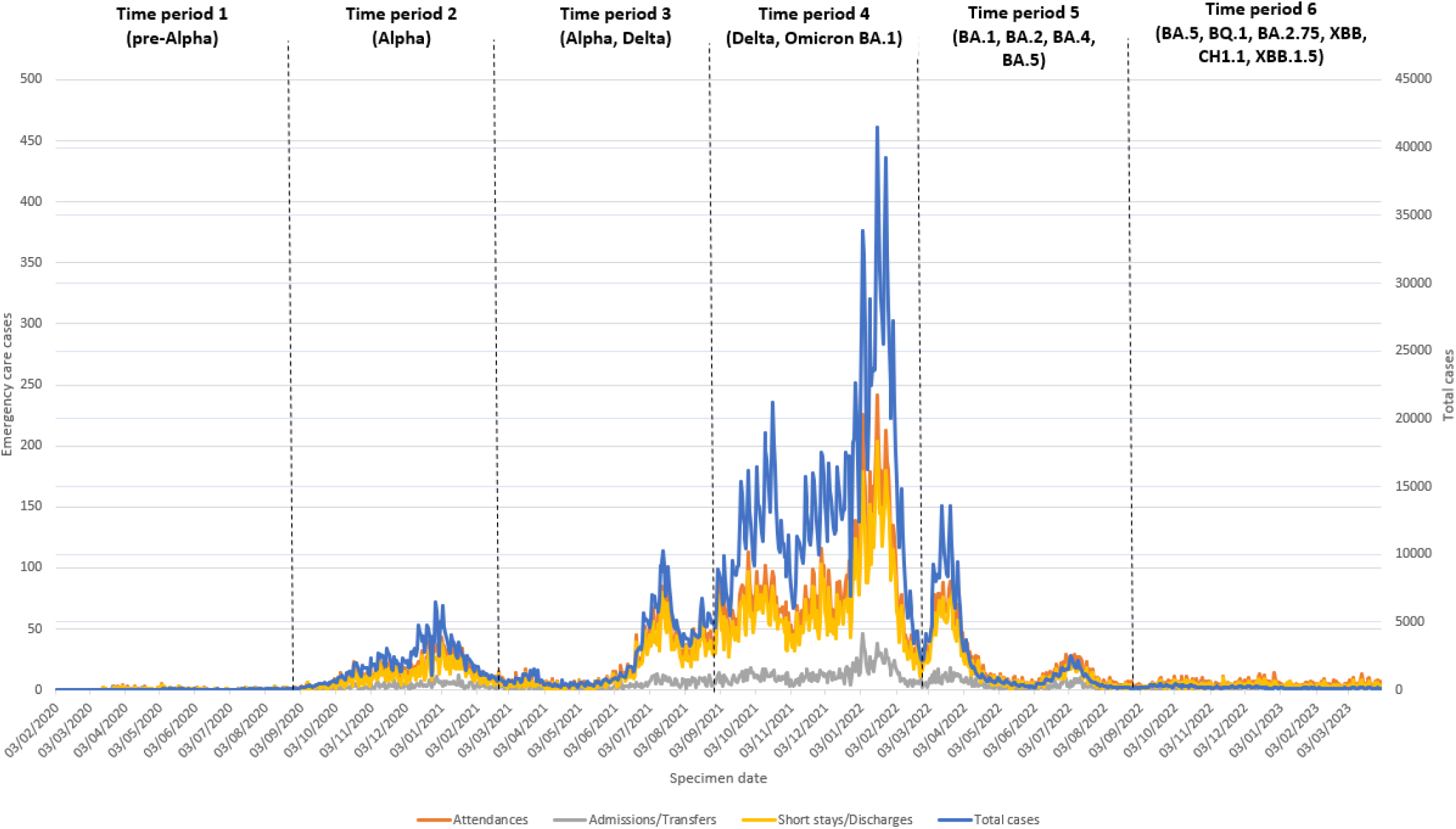
Epidemiological curve showing i) the total number of cases < 18 years of age testing positive for SARS-CoV-2 3rd February 2020 - 31st March 2023* (blue) ii) the number attending an emergency department (red) iii) the number admitted (grey) and iv) discharged** (yellow), by specimen date. * Labelled are the major variants associated with each time period, as per SGSS surveillance data and ONS reporting (6,34,35) ** Emergency care outcomes of attendance were grouped as admitted: admission to inpatient care, or transfers to other acute trusts; or discharged: admission to short stay admission unit (<24 hr in ED) or discharged

Regarding vaccination status, there was a higher proportion of unvaccinated CYP cases (58.7%) than there were vaccinated (41.1%). Unvaccinated individuals encompassed a comparatively greater proportion of ED attendances at 68.0%. Looking at attendance outcomes there was no significant difference in the number of admissions compared to discharges across all vaccination status groups (p = 0.062), with unvaccinated CYP making up a slightly higher proportion of admissions (69.4%) than discharges (67.7%). Further to this, a breakdown of the vaccination status of CYP attending emergency care by age can be seen in Table 2. The majority of those who were vaccinated at least once (n = 8,487) were aged 15-17 (47.3%) and 10-14 (41.4%), consistent with the order in which these age groups were included in the vaccination programme (10,11).

**Table 2.** Vaccination status of under 18 emergency care attendances by age group*.

| Age | Unvaccinated | | Vaccinated ($\geq 1$ dose) | |
| --- | --- | --- | --- | --- |
|  | n | % | n | % |
| <1 | 3,832 | 14.40 | 7 | 0.03 |
| 1-4 | 4,764 | 17.91 | 82 | 0.31 |
| 5-9 | 4,365 | 16.41 | 873 | 3.28 |
| 10-14 | 3,500 | 13.16 | 3,511 | 13.20 |
| 15-17 | 1,625 | 6.11 | 4,014 | 15.09 |
| Total | 18,086 | 67.99 | 8,487 | 31.90 |

\*The 8 CYP who died following attendance have been excluded
\*\*There were 31 unlinked records

## Conclusion

This descriptive analysis has summarised the epidemiology of CYP with SARS-CoV-2 infection attending emergency departments in England and demonstrates the utility of this dataset for supporting communicable disease surveillance. A major strength of this analysis is that it encompasses the start of the pandemic period up until changes in testing policy took place on 1^st^ April 2023; including ending PCR testing outside NHS settings, routine asymptomatic testing in care homes, and routine asymptomatic testing across all health and social care settings (12). Alongside the linkage of infection data from SGSS with emergency care and vaccination data, ECDS provides useful insight into the characteristics of CYP who attend emergency care, and their outcomes, yielding detailed data on emergency care attendance that would help describe health care use associated with infectious diseases.

Using ECDS data in conjunction with HES could add additional value and information on hospital attendances and outcomes, particularly as there has been a rise in emergency care visits in recent years among children and young people (13); children who may otherwise present to the GP with illnesses that would not usually get referred to ED for further examination, are now going straight to emergency departments to receive care. Therefore, having data on attendance to emergency care services allows us to gain fuller insights into health seeking behaviours.

There is massive value in the use of ECDS data compared to HES. HES data is not reported until after the completion of a hospital episode, so ongoing hospital admissions are not included in updates to the dataset. In comparison, the ECDS dataset is regularly updated using information reported by clinicians working in acute healthcare services who are able to input data, including recording hospital admissions after ED attendance, by the point of patient discharge from emergency care (14,15). This means that patient hospital admissions can be caught in ECDS before it becomes available in SUS. In addition, ED attendance data recorded in ECDS is already being routinely used to monitor trends for pre-defined conditions by the syndromic surveillance team at UKHSA (16,17). Therefore, by further linking ECDS data to SGSS data for infectious disease, this study shows the potential for the dataset to provide rapid insights during future infectious disease incidents. As well as demographic characteristics of CYP attending emergency services, outcomes of these attendances can inform assessments of severity during outbreaks.

However, it must be noted that use of ECDS for the surveillance of infectious diseases is restricted in a similar way to other routine datasets, in that it is reliant on testing practices and capacity. The levels of testing in the under 18 population changed throughout the study, especially following the publication of the “Living With COVID” policy in February 2022, which included the removal of free universal testing for the general public. In addition, routine testing was no longer required in education and social care settings, which will have affected the amount of testing carried out by this group post-Delta and Omicron BA.1 waves (i.e. Time Periods 5 and 6) (18,19). In our analysis, the periods with the lowest number of cases and attendances were Time Periods 1 and 6. This reflects times when testing was not as widespread, possibly meaning that not all individuals with the relevant presentations had been tested for SARS-CoV-2 infection in relation to having attended emergency care (18,20–24). However, Time Periods 1 and 6 had the highest proportions of cases attending ED. This high proportion could reflect limited community testing. Also of interest, would be the severity of variants circulating during that time as well as potential immunity waning from vaccinations and prior infections, or lack thereof (23-25). Therefore, it is difficult to know the true number of infected individuals who would have tested positive for COVID-19 among the CYP population prior to attending ED. Further analyses to include individuals who were tested on the day of ED attendance could account for the limitation in community testing, though this may mean those attending ED due to non-COVID related illness that is not injurious, are included.

Our findings highlight significant differences in all but one of the characteristics of young people admitted as inpatients or transferred to other acute trusts and those who stayed in ED for a short period or were discharged, and how this compares to all children diagnosed with COVID-19 in England. A higher proportion of CYP in the most deprived areas (IMD 1-3) and in the Black and Other ethnic groups were admitted to hospital following attendance at ED, than had a short stay in ED or were discharged; and there were higher proportions of CYP from non-White ethnic groups and from areas with higher deprivation levels attending emergency care, compared to in the CYP case population.

This is consistent with other studies, which have also highlighted that children living in more deprived areas had higher rates of hospital and ED attendance (not limited to COVID-19) (25). This may partially be due to difficulties in accessing primary care services; with people in more deprived areas more likely to attend ED with less severe illness, though it must be noted that they generally have poorer underlying health which may also play a role (25,26). Moreover, individuals from ethnic minority groups have disproportionately higher rates of ED admissions/attendances in comparison to those of White ethnicity (27). In addition, those from ethnic minority groups were more at-risk of contracting COVID-19 infection than their White counterparts during the pandemic (28,29). This in part may have been driven by existing socioeconomic inequalities which were worsened by the pandemic. For example, ethnic minorities are more likely to live in more deprived areas (30); and in overcrowded households, where household transmission of COVID is highest, due to difficulties in distancing between individuals (31,32). Furthermore, ethnic minorities are more likely than their White counterparts to have underlying conditions which may exacerbate the risk of adverse health outcomes stemming from COVID-19, such as hospitalisation, ED attendance and death (28,33). However, with regard to ED attendance, these differences in underlying health play a smaller role among younger populations (25).

Further work is needed to highlight the factors associated with ED attendances, as this analysis did not seek to ascertain specific factors associated with the outcomes of emergency department attendances because more detailed individual-level comorbidity data to identify clinically vulnerable persons was required. This descriptive analysis was intended to add further diversity to published COVID-19 epidemiology among CYP in England, as information on emergency department use following test-confirmed infection has been limited until now. In addition, this study highlights the benefits of how emergency care data accessed using ECDS can be used and supplemented with linkages to various other datasets to inform assessments of infection severity quickly and with clinical input, which can be useful for future outbreaks and pandemics.

## Ethics Approval Statement

UKHSA has legal permission, provided by Regulation 3 of The Health Service (Control of Patient Information) Regulations 2002 to process confidential patient information under Sections 3(i) (a) to (c), 3(i)(d) (i) and (ii) and 3(iii) as part of its outbreak response activities. This study falls within the research activities approved by the UKHSA Research Ethics and Governance Group.

## Data Availability Statement

The individual-level nature of the data used risks individuals being identified, or being able to self-identify, if the data are released publicly. Requests for access to these non-publicly available data should be directed to UKHSA.

## References

1. NHS Digital [Internet]. [cited 2023 Oct 2]. Emergency Care Data Set (ECDS). Available from: https://digital.nhs.uk/data-and-information/data-collections-and-data-sets/data-sets/emergency-care-data-set-ecds

2. Hospital Episode Statistics (HES) - NHS Digital [Internet]. [cited 2023 Dec 6]. Available from: https://digital.nhs.uk/data-and-information/data-tools-and-services/data-services/hospital-episode-statistics

3. Herbert A, Wijlaars L, Zylbersztejn A, Cromwell D, Hardelid P. Data Resource Profile: Hospital Episode Statistics Admitted Patient Care (HES APC). Int J Epidemiol. 2017 Aug;46(4):1093–1093i.

4. Hardy F, Heyl J, Tucker K, Hopper A, Marchã MJ, Briggs TWR, et al. Data consistency in the English Hospital Episodes Statistics database. BMJ Health Care Inform. 2022 Oct 28;29(1):e100633.

5. Clare T, Twohig KA, O’Connell AM, Dabrera G. Timeliness and completeness of laboratory-based surveillance of COVID-19 cases in England. Public Health. 2021 May;194:163–6.

6. Laboratory reporting to UKHSA: a guide for diagnostic laboratories [Internet]. UK Health Security Agency; 2023. Available from: https://assets.publishing.service.gov.uk/media/647749ce5f7bb700127fa25a/UKHSA_Laboratory_reporting_guidelines_May_2023.pdf

7. Tessier E, Edelstein M, Tsang C, Kirsebom F, Gower C, Campbell CNJ, et al. Monitoring the COVID-19 immunisation programme through a national immunisation Management system – England’s experience. Int J Med Inform. 2023 Feb;170:104974.

8. GOV.UK [Internet]. 2023 [cited 2023 Sep 8]. COVID-19 variants identified in the UK – latest updates. Available from: https://www.gov.uk/government/news/covid-19-variants-identified-in-the-uk-latest-update

9. Elliott P, Eales O, Steyn N, Tang D, Bodinier B, Wang H, et al. Twin peaks: The Omicron SARS-CoV-2 BA.1 and BA.2 epidemics in England. Science. 2022 May 24;376(6600):eabq4411.

10. GOV.UK [Internet]. [cited 2023 Sep 22]. All young people aged 16 and 17 in England to be offered vaccine by next week. Available from: https://www.gov.uk/government/news/all-young-people-aged-16-and-17-in-england-to-be-offered-vaccine-by-next-week

11. NHS England „ NHS rolls out COVID-19 jab to children aged 12 to 15 [Internet]. [cited 2023 Sep 22]. Available from: https://www.england.nhs.uk/2021/09/nhs-rolls-out-covid-19-jab-to-children-aged-12-to-15/

12. GOV.UK [Internet]. [cited 2023 Sep 22]. COVID-19 testing approach from April 2023. Available from: https://www.gov.uk/government/news/covid-19-testing-approach-from-april-2023

13. Ruzangi J, Blair M, Cecil E, Greenfield G, Bottle A, Hargreaves DS, et al. Trends in healthcare use in children aged less than 15 years: a population-based cohort study in England from 2007 to 2017. BMJ Open. 2020 May 5;10(5):e033761.

14. Emergency Care Data Set (ECDS) User Guide v4.5 [Internet]. NHS England; [cited 2023 Dec 6]. Available from: https://digital.nhs.uk/data-and-information/data-collections-and-data-sets/data-sets/emergency-care-data-set-ecds/ecds-guidance

15. Nyberg T, Twohig KA, Harris RJ, Seaman SR, Flannagan J, Allen H, et al. Risk of hospital admission for patients with SARS-CoV-2 variant B.1.1.7: cohort analysis. BMJ. 2021 Jun 15;n1412.

16. Hughes HE, Hughes TC, Morbey R, Challen K, Oliver I, Smith GE, et al. Emergency department use during COVID-19 as described by syndromic surveillance. Emerg Med J. 2020 Oct;37(10):600–4.

17. UK Health Security Agency. Emergency Department Syndromic Surveillance System Bulletin (England) - 2023 Week 52 [Internet]. 2024 [cited 2024 Feb 19]. Available from: https://assets.publishing.service.gov.uk/media/6596c5cf614fa20014f3a907/UKHSA_EDS SSBulletin2023wk52-1.pdf

18. Cabinet Office. GOV.UK. [cited 2022 Nov 29]. COVID-19 Response: Living with COVID-19. Available from: https://www.gov.uk/government/publications/covid-19-response-living-with-covid-19/covid-19-response-living-with-covid-19

19. GOV.UK [Internet]. [cited 2023 Dec 8]. Government reintroduces confirmatory PCR testing for assisted testing. Available from: https://www.gov.uk/government/news/government-reintroduces-confirmatory-pcr-testing

20. Lee ACK, English P, Pankhania B, Morling JR. Where England’s pandemic response to COVID-19 went wrong. Public Health. 2021 Mar;192:45–8.

21. NHS Test and Trace: the journey so far - The Health Foundation [Internet]. [cited 2023 Sep 22]. Available from: https://www.health.org.uk/publications/long-reads/nhs-test-and-trace-the-journey-so-far

22. COVID-19: Impact of the pandemic on healthcare delivery [Internet]. [cited 2023 Sep 22]. Available from: https://www.bma.org.uk/advice-and-support/covid-19/what-the-bma-is-doing/covid-19-impact-of-the-pandemic-on-healthcare-delivery

23. Hicks T, Winter A, Green K, Kierkegaard P, Price DA, Body R, et al. Care pathway and prioritization of rapid testing for COVID-19 in UK hospitals: a qualitative evaluation. BMC Health Services Research. 2021 May 31;21(1):532.

24. Rough E. Coronavirus: Testing for Covid-19 [Internet]. House Of Commons Library; 2020 Oct [cited 2023 Dec 5]. Report No.: CBP 8897. Available from: https://researchbriefings.files.parliament.uk/documents/CBP-8897/CBP-8897.pdf

25. Inequalities in Accident and Emergency department attendance, England - Office for National Statistics [Internet]. [cited 2024 Feb 29]. Available from: https://www.ons.gov.uk/peoplepopulationandcommunity/healthandsocialcare/healthcaresystem/articles/inequalitiesinaccidentandemergencydepartmentattendanceengland/march2021tomarch2022

26. Kossarova, L, Cheung R, Hargreaves D, Keeble E. Admissions of inequality: emergency hospital use for children and young people [Internet]. Nuffield Trust; 2017 Dec [cited 2024 Feb 29]. Available from: https://www.nuffieldtrust.org.uk/sites/default/files/2017-12/nt-admissions-of-inequality-web.pdf

27. Wan YI, Robbins AJ, Apea VJ, Orkin CM, Pearse RM, Puthucheary ZA, et al. Ethnicity and acute hospital admissions: Multi-center analysis of routine hospital data. eClinicalMedicine [Internet]. 2021 Sep 1 [cited 2024 Feb 29];39. Available from: https://www.thelancet.com/journals/eclinm/article/PIIS2589-5370(21)00357-6/fulltext

28. Irizar P, Pan D, Kapadia D, Bécares L, Sze S, Taylor H, et al. Ethnic inequalities in COVID-19 infection, hospitalisation, intensive care admission, and death: a global systematic review and meta-analysis of over 200 million study participants. eClinicalMedicine [Internet]. 2023 Mar 1 [cited 2024 Mar 22];57. Available from: https://www.thelancet.com/journals/eclinm/article/PIIS2589-5370(23)00054-8/fulltext#secsectitle0125

29. Mathur R, Rentsch CT, Morton CE, Hulme WJ, Schultze A, MacKenna B, et al. Ethnic differences in SARS-CoV-2 infection and COVID-19-related hospitalisation, intensive care unit admission, and death in 17 million adults in England: an observational cohort study using the OpenSAFELY platform. The Lancet. 2021 May 8;397(10286):1711–24.

30. People living in deprived neighbourhoods [Internet]. 2020 [cited 2024 Feb 29]. Available from: https://www.ethnicity-facts-figures.service.gov.uk/uk-population-by-ethnicity/demographics/people-living-in-deprived-neighbourhoods/latest/

31. Hall JA, Harris RJ, Zaidi A, Woodhall SC, Dabrera G, Dunbar JK. HOSTED— England’s Household Transmission Evaluation Dataset: preliminary findings from a novel passive surveillance system of COVID-19. International Journal of Epidemiology. 2021 Apr 9;dyab057.

32. Overcrowded households [Internet]. 2023 [cited 2024 Mar 22]. Available from: https://www.ethnicity-facts-figures.service.gov.uk/housing/housing-conditions/overcrowded-households/latest/#by-ethnicity-and-area

33. Oskrochi Y, Jeraj S, Aldridge R, Butt J, Miller A. Not by choice – the unequal impact of the COVID-19 pandemic on disempowered ethnic minority and migrant communities [Internet]. Race Equality Foundation; 2023 Jul [cited 2024 Feb 29]. Available from: https://www.doctorsoftheworld.org.uk/wp-content/uploads/2018/11/Not-by-choice.pdf

34. Coronavirus (COVID-19) latest insights - Office for National Statistics [Internet]. [cited 2024 Feb 19]. Available from: https://www.ons.gov.uk/peoplepopulationandcommunity/healthandsocialcare/conditionsanddiseases/articles/coronaviruscovid19latestinsights/infections

35. Coronavirus (COVID-19) Infection Survey, UK - Office for National Statistics [Internet]. [cited 2024 Feb 20]. Available from: https://www.ons.gov.uk/peoplepopulationandcommunity/healthandsocialcare/conditionsanddiseases/bulletins/coronaviruscovid19infectionsurveypilot/3march2023

